# Targeting Frontostriatal Reward System Connectivity and Anhedonia in Youth Depression using Continuous Theta Burst Stimulation Plus Positive Affect Training: Preliminary Evidence and Reward Network Engagement from an Open-Label Trial

**DOI:** 10.64898/2026.11.17.26368292

**Authors:** Helmet T. Karim, Neal D. Ryan, Neil P. Jones, Michelle Craske, Kristen L. Eckstrand, Rebecca B. Price, Kalyan Tripathy, Carly J. Lenniger, Manivel Rengasamy, Matthew Geramita, Benjamin Vanderschelden, Jihui Diaz, Zachary Brodnick, Halina Dour, Ashley Pogue, Charlotte Medich, Megan M. Julien, David Rogers, Erika E. Forbes

## Abstract

Anhedonia is a hallmark of depression, involves disrupted positive affect (PA), and has been linked to hyperconnectivity between the dorsomedial prefrontal cortex (dmPFC) and nucleus accumbens (NAcc). In previous work, a single session of continuous theta burst stimulation (cTBS) to the dmPFC reduced NAcc-dmPFC functional connectivity (FC), with corresponding improvement in PA. We investigated whether cTBS to dmPFC plus PA-enhancing training is associated with decreased anhedonia, depression, and NAcc-dmPFC FC, and whether FC change corresponds to anhedonia improvement. Fifty youth (15-25 years) with current depression received cTBS (2x/day, 10 days, personalized dmPFC target) plus intersession PA training. Resting-state fMRI and symptom severity were collected pre/post cTBS+PA. In all, 42% of participants responded, and depression, anhedonia, and anxiety improved. Participants with greater decrease in FC experienced more improvement in anhedonia, although NAcc-dmPFC FC remained unchanged on average. Responders exhibited greater improvement than non-responders in anhedonia and anxiety but not FC. Additionally, mutual information modeling revealed that those with greater similarity between the actual target network (based on electrical field modeling) and the intended reward network exhibited a greater decrease in FC. cTBS+PA shows some promise as an intervention for anhedonic depression in youth, with individual differences in FC change relevant to clinical outcomes and precise network targeting a critical issue for modulating the intended network and achieving clinical improvement.

Clinicaltrials.gov: NCT05487885

## Introduction

Anhedonia—difficulties with reward-focused motivation, anticipation, and enjoyment—is a key feature of treatment-resistant depression. A core symptom that also anchors a subtype of depression, anhedonia often precedes the emergence of serious psychopathology, increases risk for suicidality, and reduces the likelihood of responding to conventional pharmacologic or psychosocial treatments.^1,2^ Improving the currently disappointing rates of treatment success requires developing interventions that address anhedonia directly. A compelling issue is the need for anhedonia-focused treatments in youth, or the period from late adolescence to early adulthood, when anhedonia and depression tend to emerge. Early intervention could nudge people away from pathways toward chronic and even deadly illness onto pathways of healthy functioning.^2,3^

Translational neuroscience approaches to anhedonic depression represent a departure from psychiatry’s traditional focus on negative affect systems and toward addressing disruptions of reward and positive affect (PA) systems. Anhedonia’s neural mechanisms are postulated to involve neural reward systems,^4^ including regions in frontostriatal circuitry (e.g., nucleus accumbens, dorsal striatum, and dorsomedial prefrontal cortex). Based on altered function in reward systems in youth with depression and anhedonia,^5–7^ our guiding hypothesis is that youth with depression exhibit excessive dmPFC response to reward, which serves to enhance NAcc-dmPFC functional connectivity (FC), dampen ventral striatum (VS) responding, and generate anhedonia. Thus, our goal was to improve anhedonia and depression by reducing dmPFC activity through continuous theta burst stimulation (cTBS) to the frontostriatal reward system via a dmPFC target. This condensed version of transcranial magnetic stimulation (TMS) putatively inhibits cortical function, and the reward-system target holds promise for reducing anhedonia.

This study’s neuromodulation protocol is motivated by our experimental findings that a single cTBS session to dmPFC with individual target identification reduced NAcc-dmPFC FC in youth with depression, which was associated with greater increases in PA.^8^ Not only does cTBS to dmPFC have potential as a treatment approach, but clinical improvement could depend on individual differences in response of FC to TMS, with people whose symptoms decrease also being those whose network function changes as intended. In addition to condensed schedules, individualized targeting, and focus on reward systems, another issue deserving attention in TMS is the use of complementary psychosocial approaches to create an optimal context for modulation. For anhedonia and neuromodulation of the reward system, this could include augmenting cTBS to dmPFC with cognitive-behavioral activities aimed at increasing PA. PA Treatment (PAT)^9^ is an innovative cognitive-behavioral intervention for depression with efficacy in addressing anhedonia and reward systems specifically (while also reducing negative affect).^10,11^ Finally, while neuronavigation based on individual functional mapping within a target region or network is thought to optimize the effects of TMS, an important consideration is whether the network stimulated by TMS matches the intended network—that is, how accurately stimulation targets the intended network. Electrical field (E-field) modeling combined with resting state FC can address this question.

Our primary question was whether, in youth depression, reward-system FC and anhedonia improve with cTBS to dmPFC plus PA training. This open-label trial applied a 10-day, 20-session cTBS protocol, with the interval between each visit’s two cTBS sessions used to conduct PA training, operationalized as brief cognitive and behavioral exercises adapted from PAT. Rather than focus on depression as the primary outcome, we sought to investigate associations with change in anhedonia, given the treatment’s goal of targeting its neural mechanisms. We secondarily examined whether changes in anhedonia and FC were intercorrelated and whether treatment response, defined as reduction in depression severity, was related to change in FC or anhedonia. Finally, to consider a potentially influential source of variability in clinical and neural change, we investigated whether accuracy in engaging the reward network was related to change in FC. There were several innovative elements: We used cTBS, which can be delivered in less than 2 minutes; combined cTBS with PA training; developed a 2-week, 20-session protocol; and individually targeted dorsomedial PFC. Given these elements, as well as the preliminary nature of this work, we focused on a single neural target with an open-label design (without sham control). The study was conducted in youth, given the value of intervention early in clinical course and brain development and based on findings that TMS is safe and well tolerated in adolescents.^12^ cTBS has further advantages in this population because of its shorter duration and lower-intensity stimulation.^13^

## Materials and Methods

### Participants

We enrolled 123 participants age 15-25 years (78% female, 64% White) in a longitudinal study of anhedonic depression with optional cTBS; 50 chose at entry to receive cTBS (Figure 1) and 47 completed (1 withdrew, 2 were lost to follow-up). Demographic and clinical characteristics of participants are reported in Table 1.

**Figure 1.**
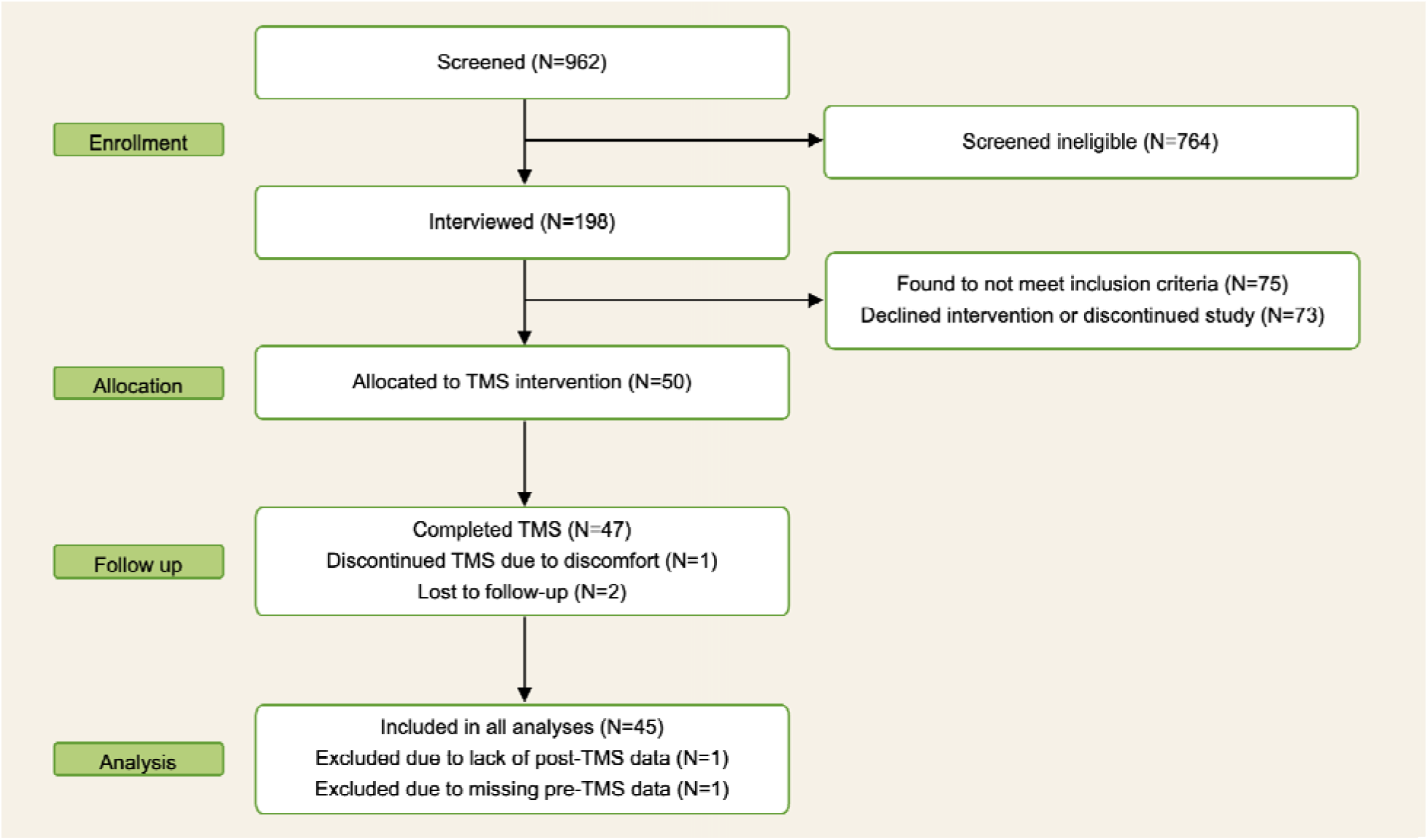
CONSORT flow diagram for open-label study of cTBS+PA training targeting frontostriatal reward system in youth depression and anhedonia (TMS: transcranial magnetic stimulation)

**Table 1.**
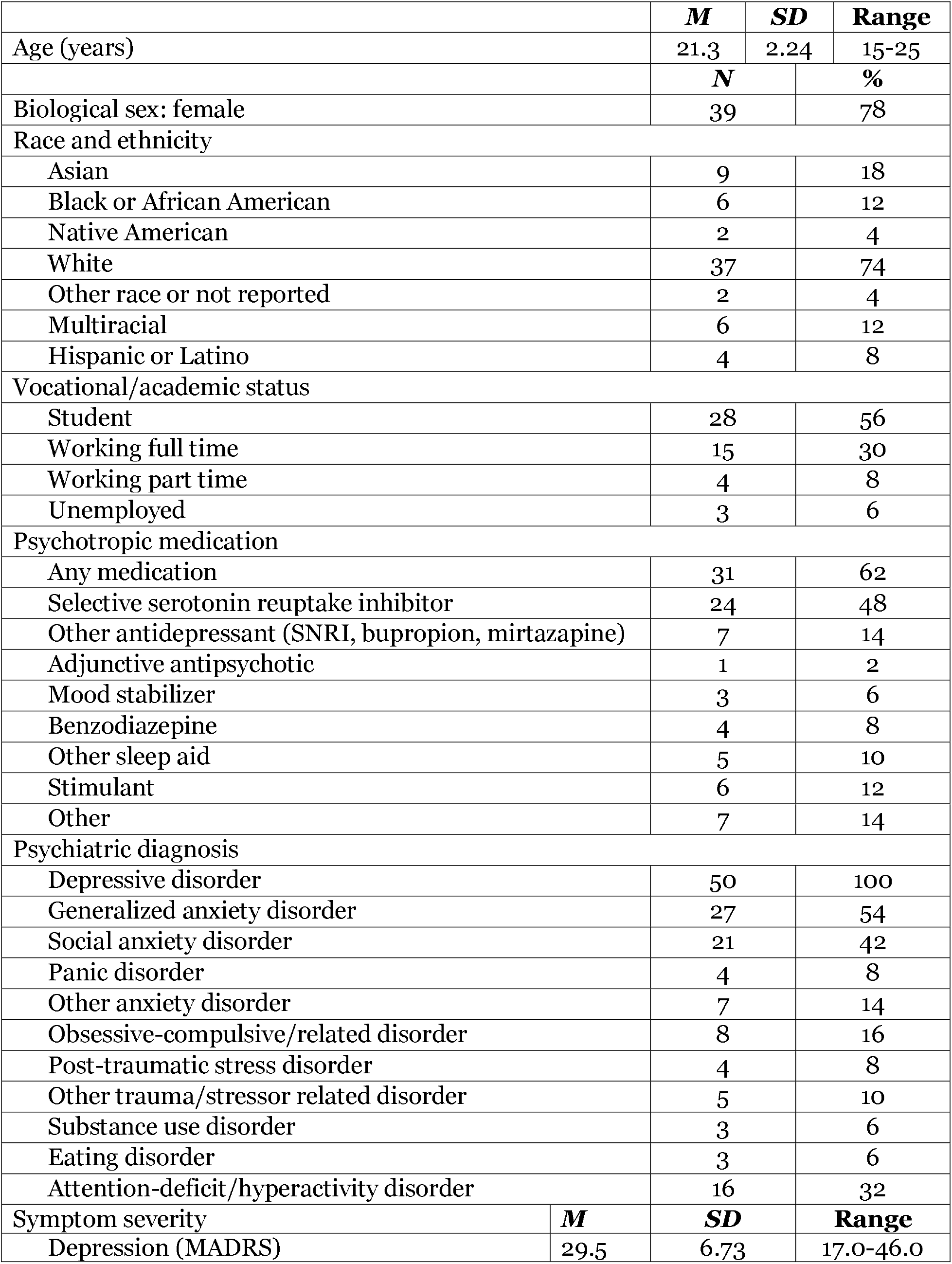
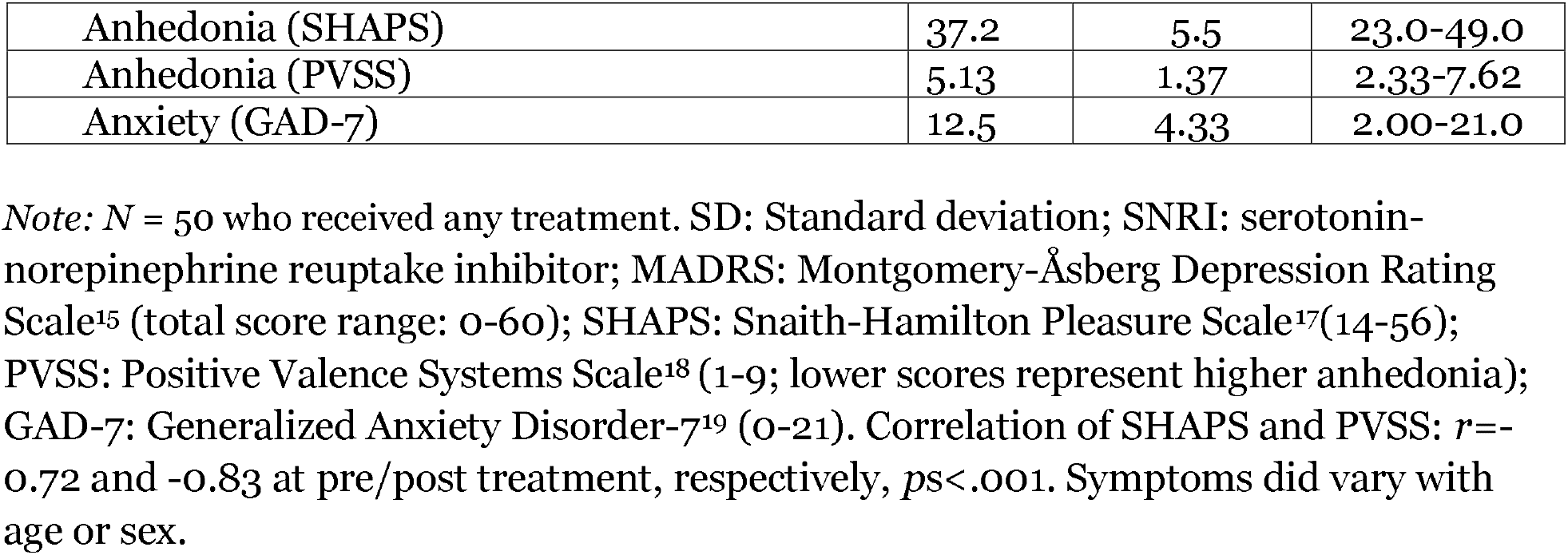
Demographic and clinical characteristics of participants.

|  | ***M*** | ***SD*** | **Range** |
|---|---|---|---|
| Age (years) | 21.3 | 2.24 | 15-25 |
|  | ***N*** | **%** |  |
| Biological sex: female | 39 | 78 |  |
| Race and ethnicity |  |  |  |
| Asian | 9 | 18 |  |
| Black or African American | 6 | 12 |  |
| Native American | 2 | 4 |  |
| White | 37 | 74 |  |
| Other race or not reported | 2 | 4 |  |
| Multiracial | 6 | 12 |  |
| Hispanic or Latino | 4 | 8 |  |
| Vocational/academic status |  |  |  |
| Student | 28 | 56 |  |
| Working full time | 15 | 30 |  |
| Working part time | 4 | 8 |  |
| Unemployed | 3 | 6 |  |
| Psychotropic medication |  |  |  |
| Any medication | 31 | 62 |  |
| Selective serotonin reuptake inhibitor | 24 | 48 |  |
| Other antidepressant (SNRI, bupropion, mirtazapine) | 7 | 14 |  |
| Adjunctive antipsychotic | 1 | 2 |  |
| Mood stabilizer | 3 | 6 |  |
| Benzodiazepine | 4 | 8 |  |
| Other sleep aid | 5 | 10 |  |
| Stimulant | 6 | 12 |  |
| Other | 7 | 14 |  |
| Psychiatric diagnosis |  |  |  |
| Depressive disorder | 50 | 100 |  |
| Generalized anxiety disorder | 27 | 54 |  |
| Social anxiety disorder | 21 | 42 |  |
| Panic disorder | 4 | 8 |  |
| Other anxiety disorder | 7 | 14 |  |
| Obsessive-compulsive/related disorder | 8 | 16 |  |
| Post-traumatic stress disorder | 4 | 8 |  |
| Other trauma/stressor related disorder | 5 | 10 |  |
| Substance use disorder | 3 | 6 |  |
| Eating disorder | 3 | 6 |  |
| Attention-deficit/hyperactivity disorder | 16 | 32 |  |
| Symptom severity | ***M*** | ***SD*** | **Range** |
| Depression (MADRS) | 29.5 | 6.73 | 17.0-46.0 |
| Anhedonia (SHAPS) | 37.2 | 5.5 | 23.0-49.0 |
| Anhedonia (PVSS) | 5.13 | 1.37 | 2.33-7.62 |
| Anxiety (GAD-7) | 12.5 | 4.33 | 2.00-21.0 |

*Note:* *N* = 50 who received any treatment. SD: Standard deviation; SNRI: serotonin-norepinephrine reuptake inhibitor; MADRS: Montgomery-Åsberg Depression Rating Scale<sup>15</sup> (total score range: 0-60); SHAPS: Snaith-Hamilton Pleasure Scale<sup>17</sup>(14-56); PVSS: Positive Valence Systems Scale<sup>18</sup> (1-9; lower scores represent higher anhedonia); GAD-7: Generalized Anxiety Disorder-7<sup>19</sup> (0-21). Correlation of SHAPS and PVSS: *r*=-0.72 and -0.83 at pre/post treatment, respectively, *p*s<.001. Symptoms did vary with age or sex.

Participants were recruited through social media advertisements, a university participant registry, and community fliers. All had a depressive disorder diagnosis per the Structured Clinical Interview for DSM-5 (SCID-5),^14^ a Montgomery-Åsberg Depression Rating Scale (MADRS)^15^ score ≥ 12, and at least one failed antidepressant trial (≥6 weeks, adequate dose). The MADRS cut-off score, reflecting mild depression, was selected to include participants with a range of severity rather than to facilitate treatment-favorable results, as remission was not the specified outcome. Exclusion criteria included lifetime psychosis or bipolar disorder; serious, unstable neurological disorder; and MRI or TMS contraindications^16^ (see Supplemental Material). The local Institutional Review Board approved all activities, and the study was registered at clinicaltrials.gov (NCT05487885). All participants completed written informed consent or assent (with parent/guardian consent if <18 years old) prior to participating.

### Study Design

Participants received cTBS plus PA training over 10 visits (2 cTBS sessions and one PA training session/visit, 5 visits/week for 2 weeks). Resting fMRI and self-reported symptoms were assessed at pre/post treatment.

### Clinical Characteristics

Participants completed the SCID-5 and MADRS, with consensus diagnosis and ratings led by a licensed psychologist or psychiatrist, and questionnaires on anhedonia (Snaith-Hamilton Pleasure Scale [SHAPS]^17^; Positive Valence Systems Scale [PVSS]^18^) and anxiety (Generalized Anxiety Disorder-7; GAD-7)^19^. Given the study’s focus on anhedonia, participants completed both SHAPS (putatively addressing general anhedonia) and PVSS (putatively addressing a broader range of facets). Participants also reported mood (Positive and Negative Affect Schedule [PANAS])^20^ and anhedonia (SHAPS) before/after each cTBS visit. One participant did not complete post-treatment assessments.

### Resting-State FC

Resting-state fMRI data were used for neuronavigation and testing hypotheses about FC change. Participants underwent eyes-open, resting-state fMRI (2 runs, 400s/run, 301 volumes/run, 13 minutes and 20 seconds) on a 3 Tesla Siemens Prisma scanner with a 32-channel head coil before and after treatment. The structural T1-weighted scan and resting-state scans used multi-echo acquisition. Preprocessing and analyses were conducted in SPM12^21^ and included normalization, slice-time correction, motion correction, tedana, co-registration, smoothing, and covariate regression (see Supplemental Materials for details on acquisition and analysis).

For NAcc-dmPFC FC, the bilateral NAcc seed region was defined using the IBASPM71 atlas within WFU PickAtlas and the dmPFC was defined as the superior medial frontal cortex in the AAL atlas.^22^ Mean resting-state NAcc timeseries were extracted, Pearson correlations were conducted with all other voxels’ timeseries, then values were extracted for average NAcc-dmPFC connectivity across both runs.

### cTBS

cTBS was administered with a MagVenture Cool-Bc65 coil with MagLink research software on a MagPro X100 stimulator. Resting motor threshold (rMT) was assessed visually by placing the coil over the motor cortex and delivering single pulses with adjustments to intensity until a visible hand movement was elicited (≥5/10 trials). The hand rather than toe motor region was used for rMT because it has similar cortical distance to the dmPFC target. cTBS was delivered at 110% rMT (with variance for tolerability: lowest stimulation was 108% rMT), with the coil handle pointed toward the vertex.

The 2 daily sessions had a 1-hour inter-session interval. Pulses were applied in a theta burst pattern (30-Hz triplet bursts at 6 Hz) for one continuous train of 33.3s, a 600 pulses/session, and 12,000 pulses total.^23^ This somewhat conservative dose reflected a balance between applying a novel target and type of TMS in a young population and providing cTBS on an accelerated protocol. For personalized targeting, the NAcc connectivity map from the first resting-state scan was masked with the dmPFC region and inverse-normalized into native MPRAGE space. This functional map was loaded into neuronavigation software (Localite, Bonn, Germany), with the stimulation target for each participant defined as the dmPFC location with maximum NAcc FC. Adherence to the 2-week schedule was good, participants were generally satisfied, and 59.8% considered their mood and energy levels improved (Table 2). There were no serious adverse events. The single cTBS-related adverse event involved scalp discomfort after the motor threshold procedure.

**Table 2.** cTBS procedures: symptom severity pre/post treatment, timing and dose, and participant experiences.

| Symptom | Pre-Treatment (*N=49*) |  | Post-Treatment (*N=46*) |  | Pre-Post Change (*N=45*) |  |  |
|---|---|---|---|---|---|---|---|
|  | Mean | SD | Mean | SD | *F* | *p* | sr<sup>2</sup> |
| Depression (MADRS) | 29.5 | 6.73 | 20.7 | 7.38 | 37.3 | <0.001 | 0.26 |
| Anhedonia (SHAPS) | 37.2 | 5.5 | 32.1 | 6.79 | 9.4 | <0.005 | 0.05 |
| Anhedonia (PVSS) | 5.13 | 1.37 | 5.37 | 1.21 | 2.2 | 0.14 | 0.01 |
| Anxiety (GAD-7) | 12.5 | 4.33 | 10.1 | 5.17 | 12.2 | <0.005 | 0.05 |
| Timing and Dose |  |  |  |  |  | Mean | SD |
| Visits completed |  |  |  |  |  | 9.62 | 1.55 |
| Sessions completed |  |  |  |  |  | 19.24 | 3.10 |
| Days taken to complete TMS visits |  |  |  |  |  | 13 | 1.78 |
| Motor Threshold (% MSO) |  |  |  |  |  | 53.6 | 8.88 |
| Participant Experiences |  |  |  | Pre-Session |  | Post-Session |  |
|  |  |  |  | Mean | SD | Mean | SD |
| Pain (0-10) |  |  |  | 0.61 | 1.25 | 0.63 | 1.24 |
|  |  |  |  | N | % | N | % |
| Lightheadedness (yes/no, sessions) |  |  |  | 2 | 0.43 | 1 | 0.22 |
| Muscle Tension (yes/no, sessions) |  |  |  | 24 | 5.22 | 24 | 5.37 |
| Headache (yes/no, sessions) |  |  |  | 25 | 5.43 | 29 | 6.49 |
| Post-treatment acceptability (0-100) |  |  |  |  |  | Mean | SD |
| *I am glad I completed this procedure* |  |  |  |  |  | 72.2 | 18.4 |
| *I felt frustrated by this procedure* |  |  |  |  |  | 17.0 | 17.2 |
| *Any side effects I experienced were manageable* |  |  |  |  |  | 77.9 | 19.6 |
| *I think this procedure is effective for helping my mood and energy* |  |  |  |  |  | 59.8 | 19.5 |
| *I was satisfied by my experience with the procedure* |  |  |  |  |  | 73.0 | 19.1 |

*Note:*
(a) cTBS: continuous theta burst stimulation; MSO: maximum stimulator output; MADRS: Montgomery-Åsberg Depression Rating Scale<sup>15</sup> (0-60); SHAPS: Snaith-Hamilton Pleasure Scale<sup>17</sup> (14-56); PVSS: Positive Valence Systems Scale<sup>18</sup> (1-9; higher scores represent lower anhedonia); GAD-7: Generalized Anxiety Disorder-7<sup>19</sup> (0-21); sr<sup>2</sup>: squared semi-partial correlation effect size (0.01=small, 0.09=medium, 0.25=large).
(b) cTBS protocol included with 2 sessions per visit and 10 visits over 2 weeks.
(c) 50 individuals started treatment, 47 completed. One completer did not provide pre-treatment questionnaire data, and one did not provide post-treatment data.
(d) Repeated-measures ANOVA with pain ratings indicated no significant main effects of session number or pre/post- session difference. A timepoint × session interaction (*F*(9,387)=2.66, *p*=0.005) suggested a minor increase in pain after early but not later sessions. Logistic regression revealed no significant effects of session, timepoint, or their interaction on lightheadedness, headache, or muscle tension.

### PA Training

Each cTBS visit included a 15-20-min inter-session PA training administered by trained research staff. Providing training between cTBS sessions was intended to capitalize on changes to the reward system generated by the first cTBS administration, prime the reward system (and affective state) for response to the second cTBS session, and enhance the efficiency of the visit. Three cognitive or behavioral exercises designed to enhance PA were adapted from PAT with collaboration from MGC and HD, who developed PAT^9–11^: planning a positive event (promote motivation toward and experience of rewarding activities; 4 sessions), finding silver linings (train attention towards positive stimuli; 3 sessions), and imagining the positive (savor future pleasant events; 3 sessions).^10^

### Electrical Field (E-field) Modeling

In the SimNIBS toolbox 4.5.0,^24^ we used ‘charm’^25^ to segment the brain and reconstruct a tetrahedral finite element mesh, generating an E-field representing the stimulated cortical region for each participant. We used each individual set of target coordinates to estimate the electrical field over the cortex. We input the coordinates of each participant’s target region, intensity of stimulation (in A/s, converted from the 110% rMT and maximum stimulator output), TMS coil used, and skin-to-coil distance (approximately 2mm), and orientation of the coil to generate an E-field. Voxel-wise maps were threshold by 50V/m to estimate a region of interest as the “actual target” (50V/m is considered therapeutically active^26^). We used this threshold as TMS activates a wide region and limiting it further would suggest a focality that does not necessarily accurately represent the stimulation.

### Actual Target Network (ATN)

Between-person variability in dmPFC target locations, combined with a fairly large E-field, could lead to inadvertently modulating a network other than the intended network. To address this, we created an index for each participant to reflect the “target congruence,” or the overlap of the *intended* target network (NAcc as seed) and the *actual* target network (ATN; stimulated dmPFC area as seed). For intended target network, we used each participant’s NAcc FC map. For ATN, we modeled each participant’s voxel-wise E-field, with 50V/m threshold, and computed its whole-brain connectivity. *Mutual information* between the intended network and ATN was computed to reflect similarity or overlap of the two networks. This approach, which has been used for alignment overlap in clinical imaging, is superior to spatial correlation (e.g., Pearson statistic) for assessing networks’ similarity, as it quantifies shared information between the voxel-wise distributions and captures both linear and non-linear dependencies, rather than simply computing the correspondence of signal intensities across brain maps and assuming a linear relationship (examples in Figure 3).^27^

### Statistical Analysis

Analyses in R v4.4.3 used anhedonia (PVSS and SHAPS) and depression (MADRS) as primary clinical outcomes and anxiety (GAD7) as secondary outcome. Linear mixed effects models examined changes in clinical and FC outcomes from pre- to post-treatment, with visit as a fixed effect with unstructured correlation, subject as a random effect, and age, sex, and scanner as covariates. Exploratory analyses examined PVSS domains as outcomes (satiation, reward anticipation, reward expectancy, reward responsiveness, reward valuation, and effort valuation).

To test associations between clinical improvement and FC change, linear regressions were conducted to predict post-treatment clinical severity from change in NAcc-dmPFC FC, adjusting for pre-treatment clinical severity, age, sex, and scanner. As above, exploratory analyses were conducted with similar models, instead using PVSS subdomains. For all findings, sensitivity analyses tested whether adjusting for changes in anxiety or depression/anhedonia altered associations.

Models with mutual information between intended network and ATN tested its association with (1) NAcc-dmPFC FC change, adjusting for age, sex, and scanner; and (2) post-treatment clinical severity, adjusting for pre-treatment severity, age, sex, and scanner.

To examine differences between treatment responders (defined as ≥30% improvement in MADRS score) and non-responders, regressions tested whether responder group predicted post-treatment anhedonia, anxiety, or FC, while adjusting for age, sex, scanner, and pre-treatment severity (for symptoms). The ≥30% criterion corresponds to what is termed “partial response” in depression treatment and to a Clinical Global Impressions-Improvement^28^ score termed “minimally improved”.^29^ This magnitude reflects some improvement and could still be considered meaningful to patients when compared with pre-treatment state or with response to other treatments. Finally, to examine changes in daily anhedonia and mood during treatment, we conducted a linear mixed effects model with day as a fixed effect and age, sex, and scanner as covariates.

## Results

In all, 21 participants were responders: 42% of all enrolled and 45.7% of those who completed treatment.

On average, anhedonia as measured by SHAPS improved with treatment [*F*(1,43.7)=9.4, *p*<0.005, sr^2^=0.05], whereas anhedonia as measured by PVSS did not [F(1,43.1)=2.2, *p*=0.14, sr^2^=0.01] (Figure 2). Depression [*F*(1,44.5)=37.2, *p*<0.001, sr^2^=0.25] and anxiety improved [*F*(1,43.6)=12.2, *p*<0.005, sr^2^=0.05]. NAcc-dmPFC FC did not change with treatment [*F*(1,45.9)=0.1, *p*=0.75869, sr^2^=0.00]. No PVSS factors changed with treatment (R^2^=0.00-0.01, *p*=0.10-0.46).

**Figure 2.**
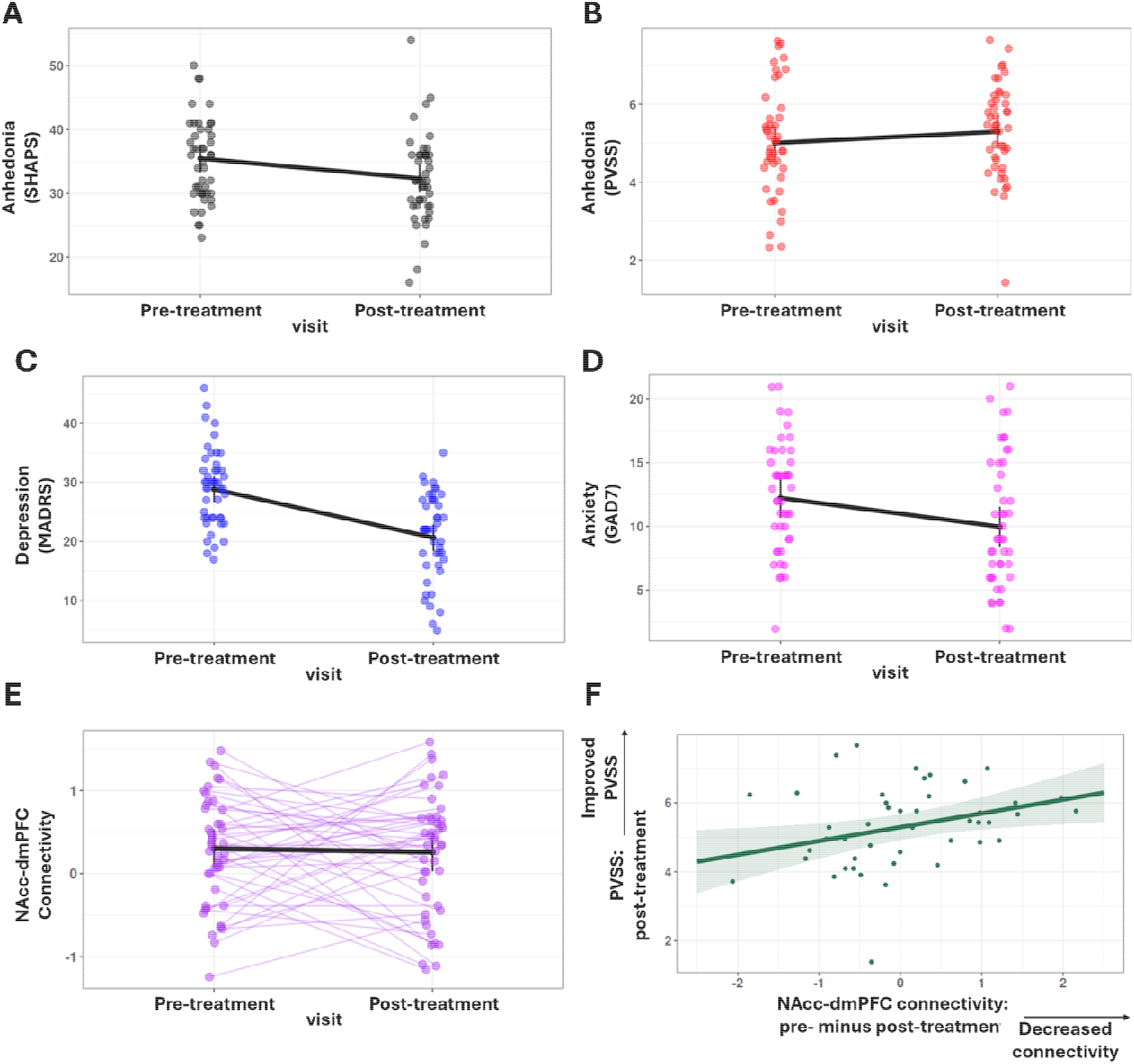
Pre- to post-treatment differences in clinical and functional connectivity (FC) measures for participants who received treatment. (A) Anhedonia - SHAPS^17^; (B) Anhedonia - PVSS^18^; (C) Depression - MADRS^15^; (D) Anxiety - GAD7^19^; (E) NAcc-dmPFC FC; and (F) improvement in post-treatment anhedonia (PVSS) vs change in NAcc-dmPFC FC. All symptom types decreased except PVSS-rated anhedonia. NAcc-dmPFC functional connectivity did not change on average. Note: for PVSS, high scores = less severity.

**Figure 3.**
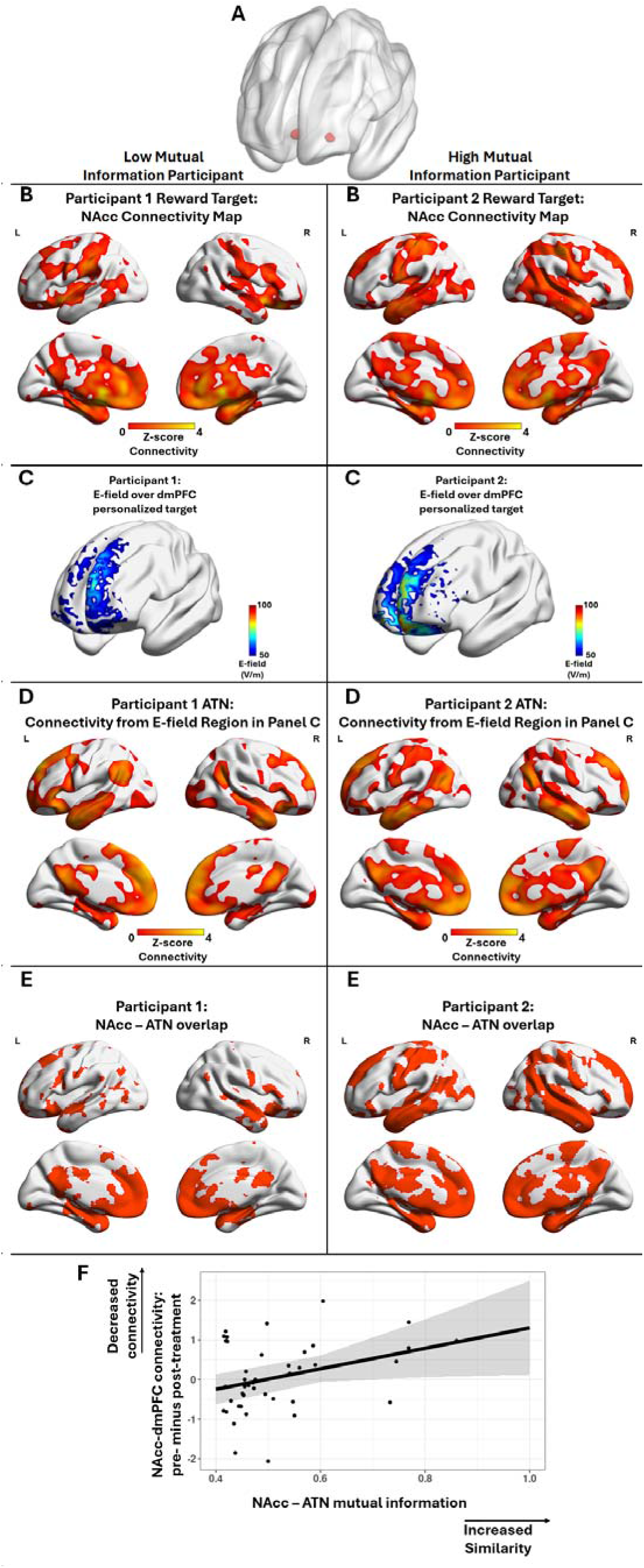
Process for generating NAcc-ATN mutual information, with examples from two participants: one with low mutual information and one with high mutual information (left and right, respectively). The intended network for cTBS stimulation (i.e., the reward network) was computed based on a NAcc seed (A) with FC to the whole brain for each participant (B). The electrical field (E-field) over the dmPFC target was modeled (C) and its whole-brain resting FC was computed (D). The E-field-derived network is defined as the Actual Target Network (ATN). The union of the NAcc network (B) with the ATN connectivity map (D) is shown in (E), indicating that the participant with high mutual information (on right) had greater overlap of the intended network (i.e., NAcc or Reward network) and ATN than the one with low mutual information (on left). Mutual information is a non-linear measure of similarity, applied in engineering approaches to medical imaging, computed here between the NAcc network and ATN. We explored whether greater NAcc-ATN mutual information was related to a larger decrease in NAcc-dmPFC connectivity from pre-post treatment (F). Participants whose target networks corresponded to or were more similar to their frontostriatal reward networks (i.e., who had higher mutual information values) showed greater improvement in functional connectivity with treatment. This highlights the value of precise targeting not just of regions but of networks when personalizing cTBS to elicit optimal treatment response.

Compared with non-responders, treatment responders experienced greater improvement in anhedonia as measured by SHAPS [B=4.16 (SD=1.83), ß=0.30, *t*=2.3, *p*<0.05] but not as measured by PVSS [B=-0.35 (SD=0.33), ß=-0.15, t=-1.1, *p*=0.30]; greater improvement in anxiety [B=2.86 (SD=1.28), ß=0.28, t=2.3, p<0.05]; and no differences in FC [B=0.02 (SD=0.25), ß=0.01, *t*=0.07, *p*=0.95].

In tests of association between change in FC and clinical outcome (Figure 2), decreases in NAcc-dmPFC FC were associated with improvement in anhedonia (increased PVSS) [B=0.41 (SD=0.16), ß=0.33, *t*=2.6, *p*<0.05]. Adjusting for changes in anxiety and depression did not alter this association [B=0.40 (SD=0.16), ß=0.32, t=2.5, p<0.05]. In exploratory analyses, this change in connectivity was primarily driven by anhedonia subdomains: PVSS reward valuation [B=1.59 (SD=0.50), ß=0.37, t=3.2, p<0.005], reward anticipation [B=1.76 (SD=0.10), ß=0.39, *t*=3.0, *p*<0.005], and reward responsiveness [B=1.76 (SD=0.81), ß=0.28, *t*=2.2, *p*<0.05]. These associations did not change when adjusting for changes in anxiety and depression. Change in FC was not associated with changes in anhedonia as measured by SHAPS [B=-1.40 (SD=0.96), ß=-0.20, *t*=-1.5, *p*=0.15], depression [B=-1.47 (SD=1.19), ß=-0.19, t=-1.2, *p*=0.22], or anxiety [B=-0.49 (SD=0.67), ß=-0.09, *t*=-0.7, *p*=0.47].

An exploratory analysis indicated that greater mutual information between each participant’s intended NAcc network and ATN—that is, more precise targeting of the reward network—was associated with greater decrease in NAcc-dmPFC FC [B=2.51 (SD=1.24), ß=0.32, *t*=2.0, *p*<0.05]. Mutual information was not associated with post-treatment clinical severity on any measure.

Across the 10 treatment days, SHAPS anhedonia decreased [F(11,493)=3.8, *p*<0.001, sr^2^=0.02], PA increased [*F*(9,405)=2.0, *p*<0.05, sr^2^=0.01], and NA decreased [*F*(9,360.3)=11.3, *p*<0.001, sr^2^=0.06] (Supplemental Figure S1).

## Discussion

In the first study to target the neural reward system as a treatment for depression in youth, we examined a novel protocol of cTBS with a personalized dmPFC target twice/day plus inter-session PA training. Depression, anhedonia, and anxiety all improved pre/post treatment, and daily PA, negative affect, and anhedonia improved steadily across treatment. While mean NAcc-dmPFC FC across participants did not decrease with treatment, individual differences were important: participants with greater decrease in FC experienced greater improvement in anhedonia. Treatment responders experienced greater decreases in anhedonia and anxiety than non-responders. Lack of decrease in FC could be partly explained by participant-level misalignment between the network we intended to target (NAcc-based reward network) and the network we in fact modulated (ATN). Specifically, greater NAcc-ATN alignment was associated with a greater decrease in NAcc-dmPFC FC. Findings are consistent with our previous work, and this novel protocol shows potential as a treatment, which is critical given anhedonia’s association with treatment resistance.^3^

Unlike studies applying TMS to dmPFC to enhance emotion regulation (e.g., in borderline personality disorder^30^), our goal was to reduce NAcc-dmPFC FC based on the hypothesis that depression (and perhaps more specifically, anhedonia) involves dampening of reward sensitivity. This hypothesis was generally supported, and findings have meaning for models of reward disruption in depression.^7^ This work extends our earlier experimental, within-participant study using similar target personalization, in which cTBS but not intermittent TBS (iTBS) or sham TBS to dmPFC in youth with depression decreased NAcc-dmPFC connectivity, which was associated with increased PA.^8^ Other TMS studies of depression have targeted the dmPFC, with some evidence for treatment effects, although none have been conducted in youth, paired with PA enhancement, or focused on anhedonia. For example, a randomized controlled trial^31^ found that while depression improved with 1Hz repetitive TMS (rTMS), 10Hz rTMS (inhibitory and excitatory, respectively), or sham rTMS, neither active condition was superior to sham.^32^ In another study using deep TMS with an H7 coil (largely dmPFC target) or an H1 coil (largely lateral PFC target), both treatments were similarly associated with reductions in depression. In the rare studies investigating either anhedonia as an outcome or the association of circuit-level change with symptom reduction, anhedonia appears to decrease with iTBS to dmPFC (which putatively has opposite effects to cTBS^33^)^34^ and decreased FC between cognitive and salience regions is related to depression improvement.^35^ Thus, past and current findings highlight the value of modulating reward systems, with particular attention to anhedonia and its relation to individual differences in FC change.

As in all treatment studies, response and symptom change varied across participants. Notably, FC did not decrease on average across participants, but individual decreases were meaningful for decreases in anhedonia. Those who exhibited a reduction in FC tended to experience improvement in anhedonia, supporting our hypothesis that modulating reward-circuit function could be particularly useful for anhedonic depression. Accordingly, it was critical to examine within-person associations of FC change with anhedonia change, not simply to test the association of cTBS+PA with mean change in FC across the sample. Further, the improvement in both anhedonia and anxiety among treatment responders (i.e., those with meaningful change in depression) indicates that improvements may not be specific to anhedonia. Similarly, and as with PAT, both positive and negative affect improved with treatment.^11^ This approach could thus have value across subtypes of depression.

Because PAT improves depression, enhances positive affect, and shifts cardiac indicators of reward responsiveness in adults with depression, the PA exercises alone could have been the main driver of our observed clinical changes and brain-symptom associations.^10,11^ However, this study remains unique in applying a focused set of exercises from PAT, delivering those exercises in combination with cTBS, testing this combination in youth, and examining neural mechanisms. Future work should address the combination of cTBS and full PAT in young people and test the hypothesis that cTBS augments the small-to-medium effects of PAT on depression.

Additionally, we addressed the possibility that variability in treatment response may have been due to inadvertently targeting networks other than the reward network. We estimated each participant’s ATN and compared it to their intended target network, revealing that the expected outcome of decreased FC was greater when cTBS engaged the reward network. Still, the range of mutual information values in our sample suggests that dmPFC targeting could have been more optimal. We used the dmPFC subregion with maximum FC to NAcc as the personalized target, but we could potentially evaluate multiple regions and select the one with both maximal FC (based on fMRI) and maximal mutual information with the ATN (based on E-field modeling). TMS studies are increasingly applying E-fields to improve the precision of targeting structural regions and functional therapeutic networks,^36–39^ with the combination of E-field modeling and functional neuroimaging improving network engagement while avoiding off-target effects.^39,40^ Considering this in addition to recent findings of variability in the location and extent of brain networks in depression,^41^ targeting a personalized *region* may not be sufficient: we must locate and engage the intended network.

Given its focus on anhedonia and its facets, this study used two different anhedonia measures, yielding slightly different patterns of findings. Improvement on the SHAPS,^17^ a widely used measure in anhedonia research that is thought to assess consummatory anhedonia, was associated with treatment and was greater for responders. Improvement on the PVSS,^18^ a newer instrument designed to capture the NIMH Research Domain Criteria perspective on a broad range of PA disruptions, was associated with decrease in FC. Overall, the relation of SHAPS to treatment response could reflect the tendency for consummatory anhedonia to track with depression. Network-level change, however, could have an influence on anhedonia more generally. PVSS findings were driven by changes in valuation, anticipation, and responsiveness subdomains of the PVSS measure, where responsiveness theoretically relates most closely to the SHAPS. This difference in measures—along with their high degree of overlap yet only <70% shared variance, underscoring a differential association with outcomes—could indicate that PVSS may, as expected, better capture the broad behavioral consequences of changes in NAcc-dmPFC FC, with even modest improvements potentially driving improvements in depression and anxiety.

Clinically, these findings raise issues about whether cTBS could be offered as an option for treatment-resistant depression in youth and whether TMS in general could eventually be considered among first-line treatments for those unlikely to respond to conventional treatments. Considerable work remains to understand the predictors of treatment response and translate those into personalized protocols, but the disappointing rate of response for standard treatments creates a need to develop and tailor a wider range of treatments. As TMS is safe, acceptable, and associated with lower long-term economic impact on the healthcare system^42^, its use in young people could potentially avoid chronic impairment and enhance healthy development.

This was an open-label trial in which all participants received both cTBS and PA-enhancement training. A randomized controlled trial with longer follow-up assessment and inclusion of sham cTBS and psychosocial placebo is needed to evaluate whether treatment effects are attributable to cTBS, PA training, or the combination of the two. Notably, the overall cTBS dose was a conservative one, selected based on the innovative nature of the cTBS target and the developmental period of the participants. Participants received a total of approximately 12,000 pulses—in contrast to the standard 18,000 pulses in iTBS for depression—and results could thus reflect a more modest effect, as well as a smaller proportion of participants classified as responders. Indeed, the low rate of discomfort experienced by participants could indicate that a higher dose would be feasible. In future studies, it may be important to evaluate whether more pulses (e.g., 18,000 or 36,000) or a different schedule (e.g., 3x/day) would result in greater improvement. PA training exercises were adaptations from PAT^11^, and a more formal approach using the full treatment is warranted. We evaluated alignment of the intended and actual networks post hoc, and future studies could aim for strong alignment before starting TMS. Finally, our mostly female, relatively diverse sample reflects the regional population of young people with depression but limits our ability to evaluate differences by biological sex or to understand treatment effects in ethnic minority, underserved, and underrepresented populations.

This work represents a promising and valuable first step, generating potential hope for a future treatment that could ameliorate disruptions in PA and may be particularly effective at helping those with anhedonic depression. Such a treatment would hold special promise for youth, who are early in their clinical course, in a developmental period of neural plasticity, and building a foundation of functioning at a critical point in the lifespan.

## Acknowledgments

The authors wish to thank the participants and their families.

## Author Contributions

HTK: Conceptualization, methodology, data curation, formal analysis, writing—original draft, writing—review and editing. NDR: Conceptualization, investigation, methodology, writing—review and editing. NPJ: Conceptualization, investigation, methodology, writing—review and editing. MC: Methodology, writing—review and editing. KLE: Investigation, writing—review and editing. RBP: Conceptualization, investigation, writing—review and editing. KT: Methodology, data curation, formal analysis, writing— review and editing. CJL: Investigation, methodology, project administration, supervision, writing—review and editing. MR: Investigation, writing—review and editing. MG: Investigation, writing—review and editing. BV: Investigation, writing— review and editing. JD: Conceptualization, methodology, formal analysis, writing— review and editing. ZB: Investigation, data curation, project administration, visualization, writing—review and editing. HD: Methodology, supervision, writing— review and editing. AP: Investigation, data curation, project administration, writing— review and editing. CM: Investigation, data curation, project administration, visualization, writing—review and editing. MMJ: Investigation, data curation, project administration, writing—review and editing. DR: Investigation, data curation, project adminsitration. EEF: Conceptualization, investigation, methodology, funding acquisition, resources, supervision, data curation, formal analysis, writing—original draft, writing—review and editing.

## Funding

Work on “Anhedonic Depression in Adolescents and Young Adults: Capturing Phenotypes and Disrupting Mechanisms at a Critical Developmental Point” was supported by Wellcome Leap as part of the Multi-Channel Psych (MCPsych) Program. This work was additionally funded by R21MH117400 and R01MH127014. HTK was funded on K01MH122741.

## Competing Interests

EEF has been a member of the scientific advisory board of Aether MindTech. The other authors declare no conflicts of interest. Wellcome Leap MCPsych evaluated the manuscript prior to publication for IP considerations but did not influence any of the analyses.

## Data availability

Data are available upon approval of reasonable request to EEF.

## Notes

### Clinical Trial

NCT05487885

### Author Declarations

IRB of the University of Pittsburgh gave ethical approval for this work.

## References

1. Pizzagalli DA. Toward a Better Understanding of the Mechanisms and Pathophysiology of Anhedonia: Are We Ready for Translation? Am J Psychiatry. 2022 Jul;179(7):458–69. doi:10.1176/appi.ajp.20220423 PubMed PMID: 35775159; PubMed Central PMCID: PMC9308971.

2. McMakin DL, Olino TM, Porta G, Dietz LJ, Emslie G, Clarke G, et al. Anhedonia predicts poorer recovery among youth with selective serotonin reuptake inhibitor treatment-resistant depression. J Am Acad Child Adolesc Psychiatry. 2012 Apr;51(4):404–11. doi:10.1016/j.jaac.2012.01.011 PubMed PMID: 22449646; PubMed Central PMCID: PMC3536476.

3. Gupta T, Eckstrand KL, Forbes EE. Annual Research Review: Puberty and the development of anhedonia - considering childhood adversity and inflammation. J Child Psychol Psychiatry. 2024 Apr;65(4):459–80. doi:10.1111/jcpp.13955 PubMed PMID: 38391011; PubMed Central PMCID: PMC10939801.

4. Pizzagalli DA. Depression, Stress, and Anhedonia: Toward a Synthesis and Integrated Model. Annual Review of Clinical Psychology. 2014 Mar 28;10(Volume 10, 2014):393–423. doi:10.1146/annurev-clinpsy-050212-185606

5. Healey KL, Morgan J, Musselman SC, Olino TM, Forbes EE. Social anhedonia and medial prefrontal response to mutual liking in late adolescents. Brain and Cognition. 2014 Aug 1;Special Issue on Reward and Regulatory Processes in Adolescence89:39–50. doi:10.1016/j.bandc.2013.12.004

6. Nielson DM, Keren H, O’Callaghan G, Jackson SM, Douka I, Vidal-Ribas P, et al. Great Expectations: A Critical Review of and Suggestions for the Study of Reward Processing as a Cause and Predictor of Depression. Biological Psychiatry. 2021 Jan 15;Adolescent Brain Development and Psychopathology89(2):134–43. doi:10.1016/j.biopsych.2020.06.012

7. Forbes EE, Dahl RE. Research Review: altered reward function in adolescent depression: what, when and how? J Child Psychol Psychiatry. 2012 Jan;53(1):3–15. doi:10.1111/j.1469-7610.2011.02477.x PubMed PMID: 22117893; PubMed Central PMCID: PMC3232324.

8. Gupta T, Karim HT, Jones NP, Ferrarelli F, Nance M, Taylor SF, et al. Continuous theta burst stimulation to dorsomedial prefrontal cortex in young adults with depression: Changes in resting frontostriatal functional connectivity relevant to positive mood. Behav Res Ther. 2024 Mar;174:104493. doi:10.1016/j.brat.2024.104493 PubMed PMID: 38350221; PubMed Central PMCID: PMC10956571.

9. Craske MG, Meuret AE, Ritz T, Treanor M, Dour HJ. Treatment for Anhedonia: A Neuroscience Driven Approach. Depression and Anxiety. 2016;33(10):927–38. doi:10.1002/da.22490

10. Craske MG, Meuret AE, Echiverri-Cohen A, Rosenfield D, Ritz T. Positive affect treatment targets reward sensitivity: A randomized controlled trial. J Consult Clin Psychol. 2023 Jun;91(6):350–66. doi:10.1037/ccp0000805 PubMed PMID: 36892884; PubMed Central PMCID: PMC10213148.

11. Craske MG, Meuret AE, Ritz T, Treanor M, Dour H, Rosenfield D. Positive affect treatment for depression and anxiety: A randomized clinical trial for a core feature of anhedonia. Journal of Consulting and Clinical Psychology. 2019 May;87(5):457–71. doi:10.1037/ccp0000396

12. Wang X, Zhang Z, Li Y, Gao Y, Cao Q. Efficacy and safety of transcranial magnetic stimulation in the treatment of children, adolescents and young adults with depression: A meta-analysis of randomized controlled trials. Journal of Affective Disorders. 2026 Jan 1;392:120132. doi:10.1016/j.jad.2025.120132

13. Rajapakse T, Kirton A. Non-Invasive Brain Stimulation in Children: Applications And Future Directions. Transl Neurosci. 2013 Jun;4(2). doi:10.2478/S13380-013-0116-3 Pubmed Pmid: 24163755; Pubmed Central PMCID: Pmc3807696.

14. First M, Williams J, Karg R, Spitzer R. User’s Guide for the SCID-5-RV Structured Clinical Interview For DSM-5® Disorders - Research Version. 2015.

15. Montgomery SA, Åsberg M. A New Depression Scale Designed to be Sensitive to Change. The British Journal of Psychiatry. 1979 Apr;134(4):382–9. doi:10.1192/bjp.134.4.382

16. Rossi S, Antal A, Bestmann S, Bikson M, Brewer C, Brockmöller J, et al. Safety and recommendations for TMS use in healthy subjects and patient populations, with updates on training, ethical and regulatory issues: Expert Guidelines. Clin Neurophysiol. 2021 Jan;132(1):269–306. doi:10.1016/j.clinph.2020.10.003 PubMed PMID: 33243615; PubMed Central PMCID: PMC9094636.

17. Snaith RP, Hamilton M, Morley S, Humayan A, Hargreaves D, Trigwell P. A Scale for the Assessment of Hedonic Tone: the Snaith–Hamilton Pleasure Scale. The British Journal of Psychiatry. 1995 Jul;167(1):99–103. doi:10.1192/bjp.167.1.99

18. Khazanov GK, Ruscio AM, Forbes CN. The Positive Valence Systems Scale: Development and Validation. Assessment. 2020 Jul;27(5):1045–69. doi:10.1177/1073191119869836 PubMed PMID: 31416336.

19. Williams N. The GAD-7 questionnaire. Occup Med (Lond). 2014 Apr 1;64(3):224. doi:10.1093/occmed/kqt161

20. Watson D, Clark LA, Tellegen A. Development and validation of brief measures of positive and negative affect: the PANAS scales. J Pers Soc Psychol. 1988 Jun;54(6):1063–70. doi:10.1037//0022-3514.54.6.1063 PubMed PMID: 3397865.

21. Ashburner J, Barnes G, Chen CC, Daunizeau J, Flandin G, Friston K, et al. SPM12 Manual. Wellcome Trust Center for Neuroimaging. 2016 Jul 20.

22. Rolls ET, Huang CC, Lin CP, Feng J, Joliot M. Automated anatomical labelling atlas 3. NeuroImage. 2020 Feb 1;206:116189. doi:10.1016/j.neuroimage.2019.116189

23. Goldsworthy MR, Pitcher JB, Ridding MC. A comparison of two different continuous theta burst stimulation paradigms applied to the human primary motor cortex. Clinical Neurophysiology. 2012 Nov;123(11):2256–63. doi:10.1016/j.clinph.2012.05.001

24. Saturnino GB, Puonti O, Nielsen JD, Antonenko D, Madsen KH, Thielscher A. SimNIBS 2.1: A Comprehensive Pipeline for Individualized Electric Field Modelling for Transcranial Brain Stimulation. In: Makarov S, Horner M, Noetscher G, editors. Brain and Human Body Modeling: Computational Human Modeling at EMBC 2018 [Internet]. Cham (CH): Springer; 2019 [cited 2025 Aug 15]. Available from: http://www.ncbi.nlm.nih.gov/books/NBK549569/ PubMed PMID: 31725247.

25. Puonti O, Van Leemput K, Saturnino GB, Siebner HR, Madsen KH, Thielscher A. Accurate and robust whole-head segmentation from magnetic resonance images for individualized head modeling. Neuroimage. 2020 Oct 1;219:117044. doi:10.1016/j.neuroimage.2020.117044 PubMed PMID: 32534963; PubMed Central PMCID: PMC8048089.

26. Deng ZD, Liston C, Gunning FM, Dubin MJ, Fridgeirsson EA, Lilien J, et al. Electric Field Modeling for Transcranial Magnetic Stimulation and Electroconvulsive Therapy. In: Makarov S, Horner M, Noetscher G, editors. Brain and Human Body Modeling: Computational Human Modeling at EMBC 2018 [Internet]. Cham (CH): Springer; 2019 [cited 2025 Aug 15]. Available from: http://www.ncbi.nlm.nih.gov/books/NBK549566/ PubMed PMID: 31725245.

27. Young AL, van den Boom W, Schroeder RA, Krishnamoorthy V, Raghunathan K, Wu HT, et al. Mutual information: Measuring nonlinear dependence in longitudinal epidemiological data. PLoS One. 2023 Apr 26;18(4):e0284904. doi:10.1371/journal.pone.0284904 PubMed PMID: 37099536; PubMed Central PMCID: PMC10132663.

28. Guy W, editor. ECDEU Assessment Manual for Psychopharmacology. Rockville, MD: US Department of Heath, Education, and Welfare Public Health Service Alcohol, Drug Abuse, and Mental Health Administration; 1976.

29. Nierenberg AA, DeCecco LM. Definitions of antidepressant treatment response, remission, nonresponse, partial response, and other relevant outcomes: a focus on treatment-resistant depression. J Clin Psychiatry. 2001;62 Suppl 16:5–9. PMID: 11480882.

30. Feffer K, Lee HH, Wu W, Etkin A, Demchenko I, Cairo T, et al. Dorsomedial prefrontal rTMS for depression in borderline personality disorder: A pilot randomized crossover trial. J Affect Disord. 2022 Mar 15;301:273–80. doi:10.1016/j.jad.2021.12.038 PubMed PMID: 34942227.

31. Hariton E, Locascio JJ. Randomised controlled trials—the gold standard for effectiveness research. BJOG. 2018 Dec;125(13):1716. doi:10.1111/1471-0528.15199 PubMed PMID: 29916205; PubMed Central PMCID: PMC6235704.

32. Dunlop K, Sheen J, Schulze L, Fettes P, Mansouri F, Feffer K, et al. Dorsomedial prefrontal cortex repetitive transcranial magnetic stimulation for treatment-refractory major depressive disorder: A three-arm, blinded, randomized controlled trial. Brain Stimul. 2020;13(2):337–40. doi:10.1016/j.brs.2019.10.020 PubMed PMID: 31711880.

33. Hermiller MS, VanHaerents S, Raij T, Voss JL. Frequency-specific noninvasive modulation of memory retrieval and its relationship with hippocampal network connectivity. Hippocampus. 2019 Jul;29(7):595–609. doi:10.1002/hipo.23054 PubMed PMID: 30447076; PubMed Central PMCID: PMC6525080.

34. Bodén R, Bengtsson J, Thörnblom E, Struckmann W, Persson J. Dorsomedial prefrontal theta burst stimulation to treat anhedonia, avolition, and blunted affect in schizophrenia or depression - a randomized controlled trial. J Affect Disord. 2021 Jul 1;290:308–15. doi:10.1016/j.jad.2021.04.053 PubMed PMID: 34020205.

35. Struckmann W, Bodén R, Gingnell M, Fällmar D, Persson J. Modulation of dorsolateral prefrontal cortex functional connectivity after intermittent theta-burst stimulation in depression: Combining findings from fNIRS and fMRI. Neuroimage Clin. 2022;34:103028. doi:10.1016/j.nicl.2022.103028 PubMed PMID: 35537216; PubMed Central PMCID: PMC9118162.

36. Dannhauer M, Gomez LJ, Robins PL, Wang D, Hasan NI, Thielscher A, et al. Electric Field Modeling in Personalizing Transcranial Magnetic Stimulation Interventions. Biol Psychiatry. 2024 Mar 15;95(6):494–501. doi:10.1016/j.biopsych.2023.11.022 PubMed PMID: 38061463; PubMed Central PMCID: PMC10922371.

37. Dannhauer M, Huang Z, Beynel L, Wood E, Bukhari-Parlakturk N, Peterchev AV. TAP: targeting and analysis pipeline for optimization and verification of coil placement in transcranial magnetic stimulation. J Neural Eng. 2022 Apr 21;19(2). doi:10.1088/1741-2552/ac63a4 PubMed PMID: 35377345; PubMed Central PMCID: PMC9131512.

38. Lynch CJ, Elbau IG, Ng TH, Wolk D, Zhu S, Ayaz A, et al. Automated optimization of TMS coil placement for personalized functional network engagement. Neuron. 2022 Oct 19;110(20):3263–3277.e4. doi:10.1016/j.neuron.2022.08.012 PubMed PMID: 36113473; PubMed Central PMCID: PMC11446252.

39. Sun W, Billot A, Du J, Wei X, Lemley RA, Daneshzand M, et al. Precision Network Modeling of Transcranial Magnetic Stimulation Across Individuals Suggests Therapeutic Targets and Potential for Improvement. Hum Brain Mapp. 2025 Aug 1;46(11):e70266. doi:10.1002/hbm.70266 PubMed PMID: 40772472; PubMed Central PMCID: PMC12329576.

40. Cao Z, Xiao X, Xie C, Wei L, Yang Y, Zhu C. Personalized connectivity-based network targeting model of transcranial magnetic stimulation for treatment of psychiatric disorders: computational feasibility and reproducibility. Front Psychiatry. 2024;15:1341908. doi:10.3389/fpsyt.2024.1341908 PubMed PMID: 38419897; PubMed Central PMCID: PMC10899497.

41. Lynch CJ, Elbau IG, Ng T, Ayaz A, Zhu S, Wolk D, et al. Frontostriatal salience network expansion in individuals in depression. Nature. 2024 Sep 19;633(8030):624–33. doi:10.1038/s41586-024-07805-2

42. Rosson S, de Filippis R, Croatto G, Collantoni E, Pallottino S, Guinart D, et al. Brain stimulation and other biological non-pharmacological interventions in mental disorders: An umbrella review. Neuroscience & Biobehavioral Reviews. 2022 Aug 1;139:104743. doi:10.1016/j.neubiorev.2022.104743

