## Supplemental Methods for "Targeting Frontostriatal Reward System Connectivity and Anhedonia in Youth Depression using Continuous Theta Burst Stimulation Plus Positive Affect Training: Preliminary Evidence and Reward Network Engagement from an Open-Label Trial"

Inclusion/Exclusion Criteria: For the phenotyping phase, participant exclusion criteria were lifetime psychosis or bipolar disorder, serious unstable neurological disorder (e.g., seizure), brain injury with loss of consciousness, moderate-severe substance use disorder for last 6 months, and MR contraindications (metal in body, weight greater than 300lb, pregnancy, etc.). For the cTBS, exclusion criteria were serious unstable respiratory or cardiovascular illness, pregnancy, high blood pressure, lifetime recreational or phencyclidine (PCP) use, or lifetime history of seizure. In the time before cTBS, we also excluded those with alcohol binge in past week before cTBS (defined as 4/5 drinks for women/men, respectively, or 3/4 drinks for girls/boys (age 15-21), respectively, over a two-hour period) or current illicit stimulant use.

Resting State FC: All scans included a high-resolution, 3-dimensional MPRAGE (Magnetization-Prepared RApid Gradient Echo) with 1mm^3^ isotropic resolution, 176 slices, repetition time (TR)=1.53s, and echo time (TE)= 2.98ms, matrix size=256x256, flip angle (FA)=9deg, inversion time (TI)=0.9s, and with Generalized Autocalibrating Partially Parallel Acquisitions (GRAPPA) of 4. We collected data on resting state with eyes open and participants instructed to view a white crosshair and stay awake. The functional resting state sequence used a multi-band echoplanar imaging (transverse orientation, A-P phase direction, 60 slices, TR = 1330ms, echo times 12.6, 29.51, 46.42, 63.33 ms, matrix size=86x86, voxel size=2.0mm^3^ isotropic resolution, multi-band factor of 4, and FA=67deg). Two runs of the resting state were run, each of which was 301 volumes (400 sec each for a total of 13 minutes and 20 seconds).

We conducted segmentation into 6 tissue classes, which also generates a deformation field for normalizing into a standard anatomical space (Montreal Neurological Institute or MNI). We created an intracranial volume mask by thresholding the gray matter, white matter, and cerebrospinal fluid tissue probability maps by 0.1 and conducting image filling and image closing in MatLab. We applied this to the MPRAGE to generate a skull-stripped MPRAGE to improve coregistration with the functional images.

For functional MRI, we first conducted slice timing correction and motion correction – applying to each echo. We then coregistered the single-band reference image from the first echo to the mean functional image generated from the first echo motion correction. We then used tedana(1) to combine multiple echos optimally, which identifies TE-dependent components as blood-oxygen level dependent (BOLD) signal. We then coregistered the single-band reference image to the skull-stripped MPRAGE and applied this transformation to the other optimally combined images. We applied the deformation field to normalize these maps and conducted spatial smoothing (full-width at half-maximum of 8mm). We then performed additional preprocessing on the resting-state data via regression of 6 parameters of motion, sinusoids corresponding to frequencies outside of the band 0.08–0.15 Hz, and five principal components of white matter and cerebrospinal fluid that were all modeled jointly to avoid re-introducing noise(2).

Our primary measure was NAcc-dmPFC connectivity. We used a functionally defined bilateral NAcc region of interest using the IBASPM71 atlas within WFU PickAtlas. We extracted the mean timeseries from the NAcc and computed the Pearson correlation with all other voxels’ timeseries in the brain. We extracted the average connectivity of the superior medial frontal cortex as defined by the automated anatomical labelling atlas 3 (AAL3)(3) – this served as the primary outcome in our analyses (see statistical analysis section). The two sessions’ connectivity was averaged.

**Results.**

Supplemental Figure 1. Changes in PANAS per daily visit. Average PA and NA are shown over each visit with 95% confidence intervals.

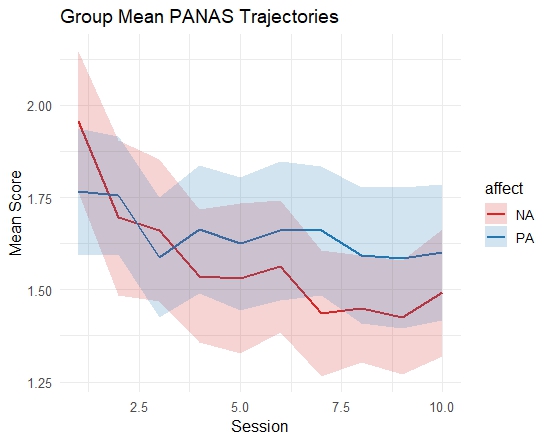

**References**

1. Kundu P, Inati SJ, Evans JW, Luh W-M, Bandettini PA. Differentiating BOLD and non-BOLD signals in fMRI time series using multi-echo EPI. Neuroimage. 2012;60:1759–1770.

2. Lindquist MA, Geuter S, Wager TD, Caffo BS. Modular preprocessing pipelines can reintroduce artifacts into fMRI data. Human brain mapping. 2019;40:2358–2376.

3. Rolls ET, Huang C-C, Lin C-P, Feng J, Joliot M. Automated anatomical labelling atlas 3. Neuroimage. 2020;206:116189.
